# Pre-pandemic activity on a myalgic encephalomyelitis/chronic fatigue syndrome support forum is highly associated with later activity on a long COVID support forum for a variety of reasons: a mixed methods study

**DOI:** 10.1101/2023.06.30.23292087

**Authors:** William U Meyerson, Rick H Hoyle

## Abstract

Encephalomyelitis/chronic fatigue syndrome (ME/CFS) and long COVID share some clinical and social characteristics. We predicted that this would lead to an increased interaction between pre-pandemic members of an ME/CFS online support community and a long COVID community. We performed a mixed-methods retrospective observational study of the Reddit activity of 7,544 users active on Reddit’s long COVID forum. From among 1600 forums, pre-pandemic activity specifically on a ME/CFS forum is the top predictor of later participation on the long COVID forum versus an acute COVID support forum. In the qualitative portion, motives for this co-participation included seeking mutual support and dual identification with both conditions. Some of this effect may be explained by pre-existing ME/CFS possibly being a risk factor for long COVID and/or SARS-CoV-2 infection being a cause of ME/CFS relapse. The high rate of ME/CFS patients seeking mutual support on a long COVID forum speaks to the longsuffering experience of these patients not feeling heard or respected, and the hope of some ME/CFS patients to gain legitimacy through the public’s growing recognition of long COVID.

## Background

Post-acute COVID-19 syndrome, better known by some as long COVID, is a recognized multisystem syndrome triggered by infection with SARS-CoV-2 that can be disabling (1). Prominent symptoms include fatigue, difficulty concentrating, changes in taste or smell, and joint or muscle pain, with different patients experiencing different subsets of symptoms (2–4). Observers have noted that prolonged fatigue, difficulty concentrating, and certain other symptoms of long COVID overlap with prominent symptoms from the multisystem disease of myalgic encephalomyelitis/chronic fatigue syndrome (ME/CFS) (5). At time of manuscript preparation, there are no known prospectively validated diagnostic tests for either ME/CFS (6) or long COVID, much less for distinguishing them from each other. Diagnosis instead is made based on the presence and duration of symptoms, exclusion of other causes, and, in the case of long COVID, guided by the timing of symptoms with respect to known or presumed SARS-CoV-2 infection (1). Commentators have long suspected that ME/CFS may likewise be triggered by viral infection, although a comprehensive causal link has not yet been definitively identified (7). This suspected viral etiology of ME/CFS also raises the possibility that SARS-CoV-2 could be one of a number of viruses with the potential to cause or worsen ME/CFS.

In addition to the at least superficial clinical similarities between long COVID and ME/CFS, there are social similarities between these two disorders. Both conditions can affect patients’ relationships and ability to work, and patients suffering from either of these disorders find that segments of society and the medical system use our ignorance about these conditions to minimize their real suffering (8,9).

Because of the social similarities and some clinical similarities between ME/CFS and long COVID, we expect that these two patient communities would interact with each other. By understanding how these two communities interact with each other, we can better learn to address the concerns of both communities.

To learn more about the interaction between ME/CFS and long COVID communities, we performed a mixed methods analysis of the interactions between support forums for these two conditions on the popular social media platform Reddit. Our use of social media as a data source is appropriate because it was in part through social media that long COVID first came to be recognized as a condition, and because social media continues to play an important role in how people get information about long COVID (10).

We predicted three kinds of involvements between pre-2020 members of Reddit’s ME/CFS support forum and Reddit’s long COVID support forum: dual identification – some users might self-identify with both conditions; mutual support – some ME/CFS-identified patients might come to the long COVID forum to seek or offer support as members of a community facing similar challenges; and professional interest in both conditions.

We first performed a case-control retrospective study to determine whether pre-pandemic activity on Reddit’s ME/CFS support forum predicted later activity on Reddit’s long COVID support forum. Then, in the qualitative portion of the study, we sought to understand the diverse motives of the users who post on both forums.

## Data and Methods

### Ethics

This study of publicly available data was determined by the Duke University Health System Institutional Review Board to not require IRB approval, with protocol number Pro00110871. The collection and analysis of data complied with the terms and conditions of Reddit data. We made no attempt to identify users nor did we purposefully recover deleted information. Per Reddit’s privacy policy, informed consent was not required for this analysis of publicly available data: “By using the Services, you are directing us to share this information publicly and freely (11).”

### Subjects

The initial case definition was the set of Reddit users with at least one post (submission or comment) on r/covidlonghaulers by April 22, 2022 and at least one post to Reddit prior to 2020. The quantitative portion of the analysis made use of two control groups. The initial “COVID+” control group was the set of Reddit users with at least one available post on the subreddit r/COVID19positive and at least one post on Reddit prior to 2020 and who were not active on r/covidlonghaulers during the study period. A second “random” control group was a random sampling of Reddit users with at least one available post after 2020 and at least one before 2020. In all groups, subjects were excluded if flagged as suspected bots (see *Bot filtration* below). The initial case and control groups were downsampled to have an equal number of users with a similar number of pre-2020 posts (see *Balancing cases and controls* below).

### Data acquisition and processing

#### Bulk download of archived data

The PushShift Reddit corpus (12), including the metadata fields of username, timestamp, and subreddit of all submissions and comments on all of Reddit from inception (June 23, 2005) through last archived date at the time (June 30, 2021), was downloaded on January 24, 2022.

#### Supplementation of bulk Reddit metadata with not-yet-archived posts

The metadata and contents of all submissions and comments on r/covidlonghaulers from inception (July 24, 2020) to the date of query (April 22, 2022) were downloaded using a custom R script that makes use of api.pushshift.io.

#### Bot filtration

Users were flagged as suspected bots if they appeared in a crowd-sourced compendium of suspected bots, botrank.pastimes.eu, which has been previously used (13) in academic work for this purpose. Users were also flagged if they contained any of following suspect, case-insensitive strings in their usernames: “bot”, “auto”, “transcribe”, “link”, “summarize”, “messenger”, “delete”, “download”, “count”, “reply”, “post”, “mention”, “metric”, “daemon”, “b0t”, “droid”, “spam”, “gif”, “click”, “detect”, or “spell”. Users flagged as suspected bots were excluded from analysis.

#### Balancing cases and controls

For each user within the cases and controls, we tabulated the number of pre-2020 submissions and comments. These counts were stratified into log2 bins, rounded down (e.g. a bin of 0 representing users with 1 pre-2020 post, a bin of 1 representing users with 2 or 3 pre-2020 posts, and so on). Users with 131,072 (2^17) or more posts were excluded. To produce the final case and control groups, users were removed through a random number generator so that there were the same number of cases, COVID+ controls, and random controls in each posting frequency bin. In almost all instances, this downsampling procedure involved removing control users since the control groups had many more users to begin with.

#### Gender assignment

Reddit users are not prompted to input their gender on their profile as they are on some other social media platforms, but some users choose to report their gender on Reddit in other ways. For example, on the r/AskMen and r/AskWomen subreddits, users are encouraged to add to their posts within the subreddit a standardized metadata tag in the form of “author flair” identifying their gender. While these metadata are sparse, their use in gender identification is an established approach in Reddit research (14). In this study, users were identified as male if they have one or more author_flair_text entries labeled with ‘♂’, ‘Male’, ‘Unverified Male’, ‘Verified Male’, ‘Male Verified’, or ‘:male: Verified!’ or one or more author_flair_css_class entries labeled with ‘male’, ‘BOY’, or ‘m’. Users were identified as female if they have one or more author_flair_text entries labeled with ‘♁’, ‘Female’, ‘Unverified Female’, ‘Verified Female’, ‘Female Verified’, or ‘:female: Verified!’ or one or more author_flair_css_class entries labeled with ‘female’, ‘GIRL’, or ‘f’. These labels were chosen after manual review for gender identification among the top 1000 most common author_flair_text values and top 1000 author_flair_css_class values, with manual rescue of gender-symmetric flair values that were not in the top 1000. For simplicity, users who identified with one gender in one post and another gender in another (<1% of gender-labeled users) were not labeled with a gender.

#### Age assignment

Reddit users are also not prompted to input their age on their profile, nor is age commonly reported with author flair, so a common approach in the literature for inferring social media user ages is through regular expressions of free text of author posts (15). The R code for the series of regular expressions for extracting age is provided in Appendix A.

#### Location assignment

A Reddit geolocation python package includes within its training data the publicly self-reported locations of more than 70,000 Reddit users (16). Countries of users were assigned for the subset of users in the cases and controls who overlapped with this training set. Given data sparsity, formal analysis of locations in cases and controls was limited to calculating the percent of labeled users who were assigned to within the United States vs outside the United States.

### Quantitative analysis

#### Calculating subreddit enrichment

We counted for each subreddit the number of unique users in the cases group, COVID+ group, and random group who had posted in that subreddit prior to 2020. We restricted analysis to the 1596 subreddits represented by at least 50 users of the cases group. For each subreddit, we calculated the odds of pre-2020 participation in that subreddit among cases relative to the most conservative control group (COVID+). Subreddits were ranked by odds ratio point estimate. Odds ratios, confidence intervals, and p-values were calculated using the epitools R package, using the default (median unbiased estimation) parameters for the confidence interval and the chi squared test for p-values; all reported p-values from this approach involved n>=5.

#### Identifying comparison medical support forums

To identify medical support forums with which to compare r/cfs, we started with the 2097 subreddits that were represented by at least 100 unique authors total between the three groups. Manual review of the names of these subreddits by a physician (WM) revealed 34 subreddits whose names were suggestive of an illness support forum. Subreddits were then excluded if 10% or more of their users among the cases were also active in r/cfs, as was the case for 17 subreddits. The remainder included multiple substance use disorder-related subreddits and other mental health-related subreddits, reflecting Reddit’s predominantly teen and young adult user base, so to minimize redundancy, only the largest represented substance use disorder-related subreddit and largest other mental health-related subreddit were included. The About panel for each included subreddit was then reviewed to confirm that the subreddits were indeed intended as illness support forums. The final list of comparison subreddits had 7 entries: r/acne, r/cancer, r/depression, r/diabetes, r/GERD, r/PCOS, and r/stopdrinking.

### Qualitative analysis

To characterize reasons why pre-2020 r/cfs users posted on r/covidlonghaulers, we used qualitative content analysis to summarize the posts of r/cfs users in r/covidlonghaulers. The first 10 submissions or comments in r/covidlonghaulers of the 160 users who posted on r/covidlonghaulers and had also posted on r/cfs prior to 2020 were read in full by WM, together with the post that these were a response to where applicable. To interpret the quantitative findings above, the team was most interested in the following themes: dual identification (users identifying with both ME/CFS and long COVID), mutual support (seeking or offering support with long COVID patients explicitly as a ME/CFS patient), and professional interest (self-identified researchers, clinicians, and venders).

Self-identification with long COVID was sufficient to count as dual identification (i.e. a pre-2020 r/cfs user who mentions on r/covidlonghaulers that he or she suffers from long COVID but does not mention ME/CFS would still count as dual identification). Implicit identification with long COVID was also coded as dual identification (e.g., “It’s been 6 months since I had COVID, when does this end?”). Self-identification with long COVID was sufficient for coding as dual identification even if the user may not have met full diagnostic criteria for long COVID (e.g. insufficient duration of symptoms). Chronic symptoms attributed by users to a COVID vaccine were not coded as dual identification. To be coded as mutual support, a user had to mention on his or her first 10 posts on r/covidlonghaulers that he or she has a chronic pre-existing illness and neither explicitly nor implicitly identify with long COVID; the user did not need to specify that the pre-existing condition was ME/CFS so long as he or she had been active on r/cfs. To be coded as professional interest, a user had to explicitly state he or she has a formal role in interacting with long COVID patients (e.g., clinician, researcher, vender) and not also mention that he or she also suffers from long COVID. All other users were coded as other/unknown.

## Results

### Sample characteristics

#### Number of posts

24,600 non-deleted submissions and 346,590 non-deleted comments were contributed by 18,132 unique users on r/covidlonghaulers from July 24, 2020 to April 22, 2022. 366 of these users, who together contributed 8,857 posts on r/covidlonghaulers, were flagged as suspected bots and removed from further analysis (see *Methods*: *Bot filtration*). 17,766 non-flagged users and their 362,333 r/covidlonghaulers posts were retained. The top 1% most prolific non-flagged users (308 or more posts on the subreddit) contributed 31.9% of all posts on r/covidlonghaulers; 75.9% of posts on the subreddit were from the top 10% most prolific users (37 or more posts on the subreddit). 5839 (32.9%) users made exactly 1 post on r/covidlonghaulers. The majority of users on the subreddit, 10,114 (56.9%), had between 2 and 36 posts. Looking more broadly to the activity of these users across all subreddits within the PushShift corpus (comprising dates June 23, 2005 to June 30, 2021), 14,423 users were included with the corpus, and these users contributed a median of 147 posts to the overall corpus. 7565 nonflagged users on r/covidlonghaulers (52.5%) had contributed at least 1 post to any Reddit forum in the PushShift corpus before 2020 and were eligible for further analysis.

#### Demographics

We next compared basic demographic information between cases and controls where available (see *Methods: Data acquisition and processing*). We found that the three groups had similar ages, gender composition, and national origin where known (see Table 1).

**Table 1:** Reddit usage and demographics of cases and controls.

|  | **longhaulers** | **COVID+** | **random** |
| --- | --- | --- | --- |
| users | 7544 | 7544 | 7544 |
| median pre-2020 posts per user<br>(interquartile range) | 134<br>(21-687.25) | 134<br>(20-683) | 133<br>(20-688) |
| median reported age<br>(interquartile range) | 30<br>(24-37) | 28<br>(23-34) | 30<br>(24.5-36) |
| percent male | 56.5% | 52.6% | 55.6% |
| percent female | 43.5% | 47.4% | 44.4% |
| percent US-based | 77.3% | 75.3% | 69.5% |
| age available | 1109 (14.7%) | 1168 (15.5%) | 1796 (23.8%) |
| gender available | 370 (4.9%) | 390 (5.2%) | 674 (8.9%) |
| country available | 66 (0.87%) | 73 (0.97%) | 177 (2.3%) |

### Subreddit enrichment

#### r/covidlonghaulers were especially likely to post on r/cfs pre-pandemic

We hypothesized that the cases (users who had posted on r/covidlonghaulers) were especially likely to have posted in r/cfs pre-pandemic as compared to controls. To test this hypothesis, we ranked each of 1596 well-represented subreddits by the number of unique users active in that subreddit pre-2020 among the cases relative to controls (see *Methods: Calculating subreddit enrichment*). The top-ranked subreddit was r/cfs. Specifically, 160 long-COVID cases had posted in r/cfs pre-pandemic, versus 27 of an equal number of equally active COVID+ users and 4 users from the random sample control group, for an odds ratio of 6.0 (95% CI 4.1-9.2, p-value 1.3 e-22 by chi squared test) versus the COVID+ group.

Secondarily, we queried the next most enriched subreddits among cases. The top-10 most enriched subreddits are shown in Table 2, which includes support forums for dysautonomia, postural orthostatic tachycardia syndrome, chronic pain, fibromyalgia, and Lyme disease (including chronic Lyme). These forums have a high rate of shared users with r/cfs. Like ME/CFS, these conditions have been described as chronic invisible illnesses (17).

**Table 2.**
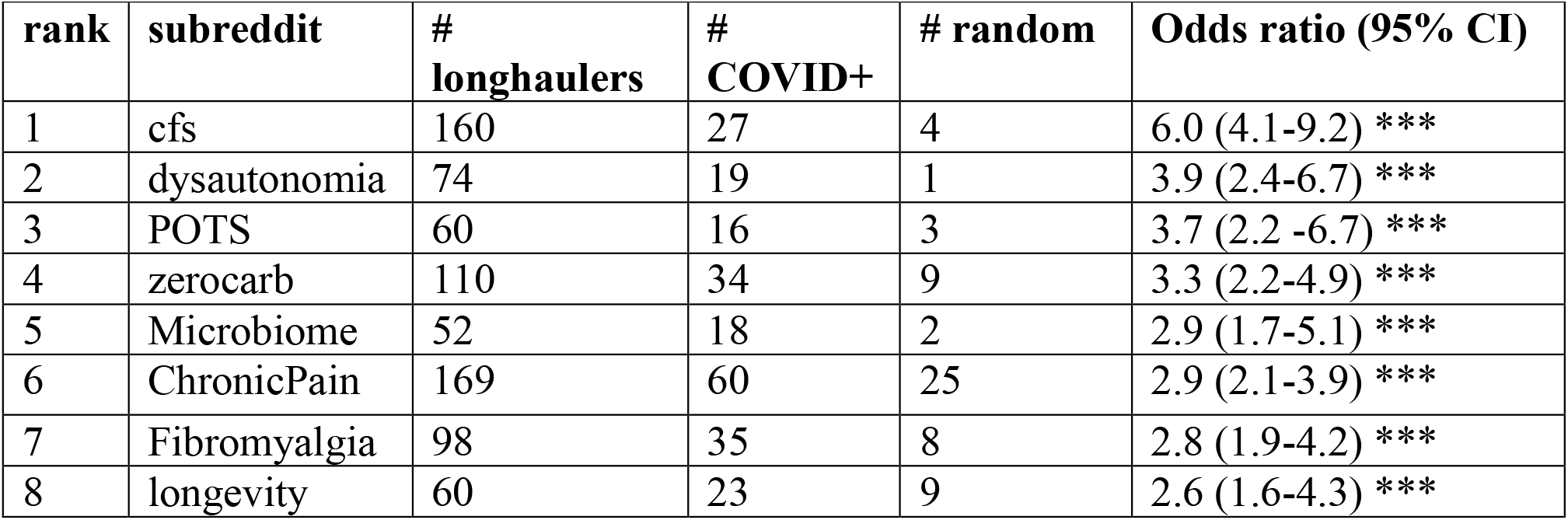
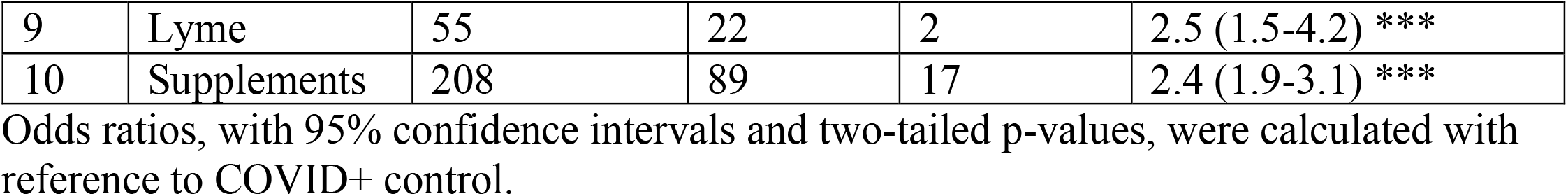
Top 10 most-enriched pre-2020 subreddits of r/covidlonghaulers.

| **rank** | **subreddit** | **#**<br>**longhaulers** | **#**<br>**COVID+** | **# random** | **Odds ratio (95% CI)** |
| --- | --- | --- | --- | --- | --- |
| 1 | cfs | 160 | 27 | 4 | 6.0 (4.1-9.2) *** |
| 2 | dysautonomia | 74 | 19 | 1 | 3.9 (2.4-6.7) *** |
| 3 | POTS | 60 | 16 | 3 | 3.7 (2.2 -6.7) *** |
| 4 | zerocarb | 110 | 34 | 9 | 3.3 (2.2-4.9) *** |
| 5 | Microbiome | 52 | 18 | 2 | 2.9 (1.7-5.1) *** |
| 6 | ChronicPain | 169 | 60 | 25 | 2.9 (2.1-3.9) *** |
| 7 | Fibromyalgia | 98 | 35 | 8 | 2.8 (1.9-4.2) *** |
| 8 | longevity | 60 | 23 | 9 | 2.6 (1.6-4.3) *** |
| 9 | Lyme | 55 | 22 | 2 | 2.5 (1.5-4.2) *** |
| 10 | Supplements | 208 | 89 | 17 | 2.4 (1.9-3.1) *** |

Odds ratios, with 95% confidence intervals and two-tailed p-values, were calculated with reference to COVID+ control.

#### Higher absolute rates of pre-2020 r/cfs posting among r/covidlonghaulers with many posts

While the relative rate of pre-2020 r/cfs posting among r/covidlonghaulers is high, the absolute percentage is modest at 2.1%. We hypothesized that this modest absolute percentage in part reflects the fact that not every Reddit user who suffers from ME/CFS or is otherwise interested in ME/CFS is active on the ME/CFS subreddit. More specifically, we hypothesized that the power to detect a pre-2020 interest in ME/CFS increases with the number of pre-2020 posts a user publishes on Reddit. To test this, we re-analyzed participation in r/cfs by cohort, this time stratifying by number of posts per author (Figure 1). We find that the proportion of users with any pre-2020 r/cfs activity substantially increases among cases but not to the same degree in controls when restricting analysis to users with many posts on Reddit overall. For example, 6.7% of cases with between 4096 (2^12) and 8191 (2^13-1) posts to Reddit overall had posted in r/cfs pre-2020, (95% CI 4.1%-10.7%), as compared with 0.79% of r/COVID19positive users (CI 0.14%-3.1%) and 0% of random users (CI 0%-1.9%) in the same posting volume interval. Moreover, there is no clear evidence of saturation, and pre-2020 activity in r/cfs among cases continues to increase with the number of pre-2020 posts over the whole range. These results suggest that the true absolute percentage of r/covidlonghaulers with a pre-2020 interest in ME/CFS is likely larger than the underpowered raw fraction of r/covidlonghaulers who posted in r/cfs pre-2020.

**Fig. 1:**
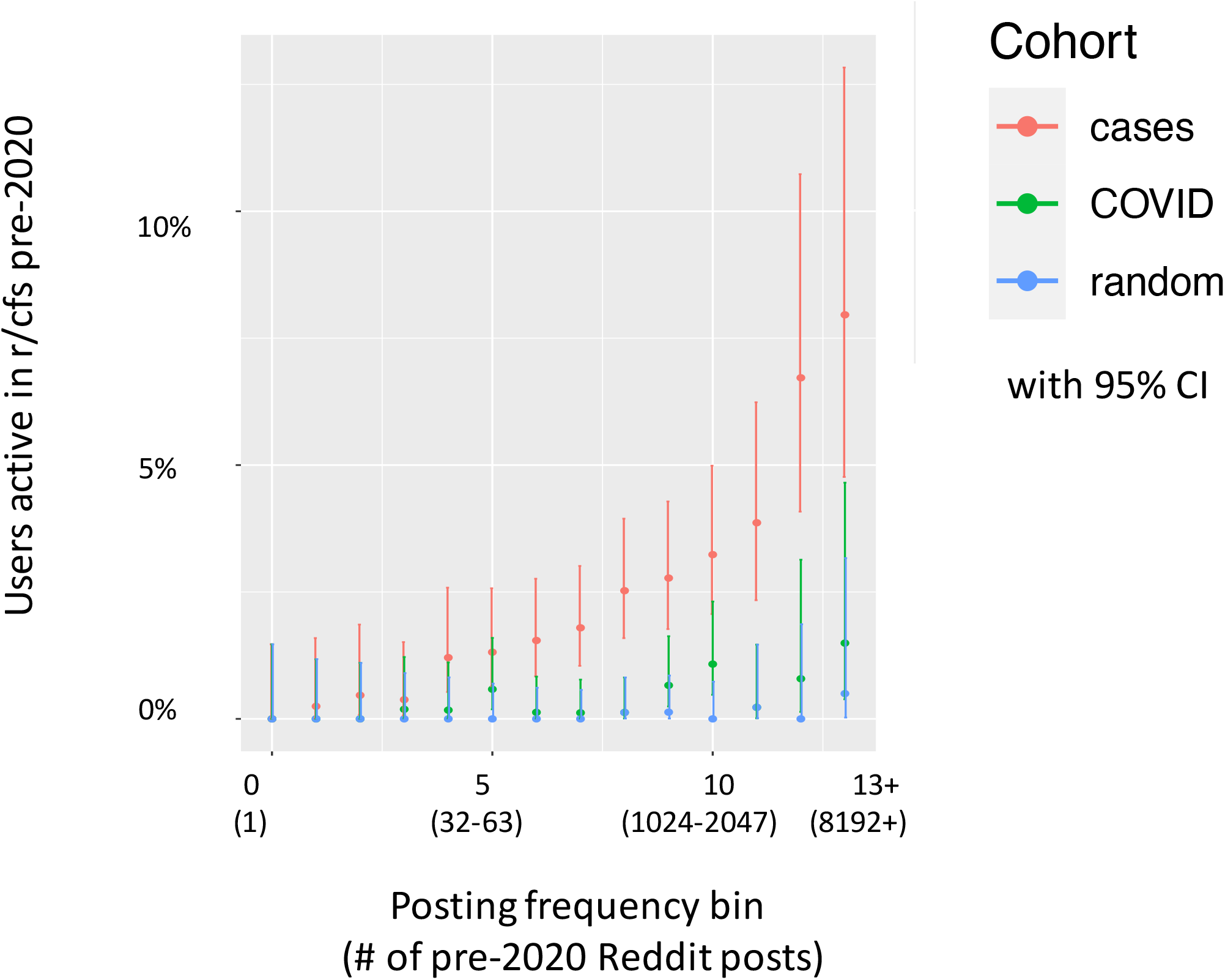
Pre-pandemic activity on r/cfs by cohort and number of posts. Y-axis represents the percentage of users within a stratum who posted at least once to Reddit’s most popular ME/CFS support forum prior to 2020. Users have been stratified by number of posts to Reddit (x-axis) and by cohort (color). Posting frequency bins per user are calculated as the log2 number of pre-2020 posts, rounded down to the nearest integer. Cohorts include cases (red): users who posted to r/covidlonghaulers, COVID (green): users who posted to r/COVID19positive, and random (blue): a random sample of recently active Reddit users. Error bars give 95% confidence intervals. Due to insufficient number of users with the highest post counts, users with 2^13 or more pre-2020 posts have been collapsed into the 13+ bin for plotting purposes.

### Does response-bias explain the r/cfs – r/covidlonghaulers overlap?

One property shared by r/cfs and r/covidlonghaulers is that the users active on these forums are interested in discussing health, including stigmatized illness, on social media. Does this explain why the users on these forums overlap? To address this question, we curated a list of support forums for other suitable illnesses to determine if these were similarly enriched among cases compared to controls (see *Methods: Identifying comparison medical support forums*). Note that this curated list excluded subreddits with a high rate of overlapping authors with r/cfs to reduce confounding, and this effectively excluded most of the top 10 enriched subreddits listed in Table 2. We found that compared to other included support forums, r/cfs is a clear outlier in terms of enrichment among cases versus controls (Figure 2). Whereas, as previously stated, pre-2020 r/cfs posting occurred with 6.0-fold higher odds among cases than in the most conservative control (COVID+), no other included support forum was comparably enriched. As an aside, it is interesting to note that the illness for the second-most enriched support forum, diabetes, was found in a recent study to have the largest point estimate for the odds ratio of developing long COVID among a set of pre-existing conditions (although the point estimate was not significant in that study) (18). These results indicate that willingness to discuss stigmatized illness online is not the main reason for the overlap between r/cfs and r/covidlonghaulers, at least in comparison to the COVID+ control. Secondarily, the COVID+ control appears to be more robust at controlling for response bias than does the random subset of active Reddit users.

**Fig. 2:**
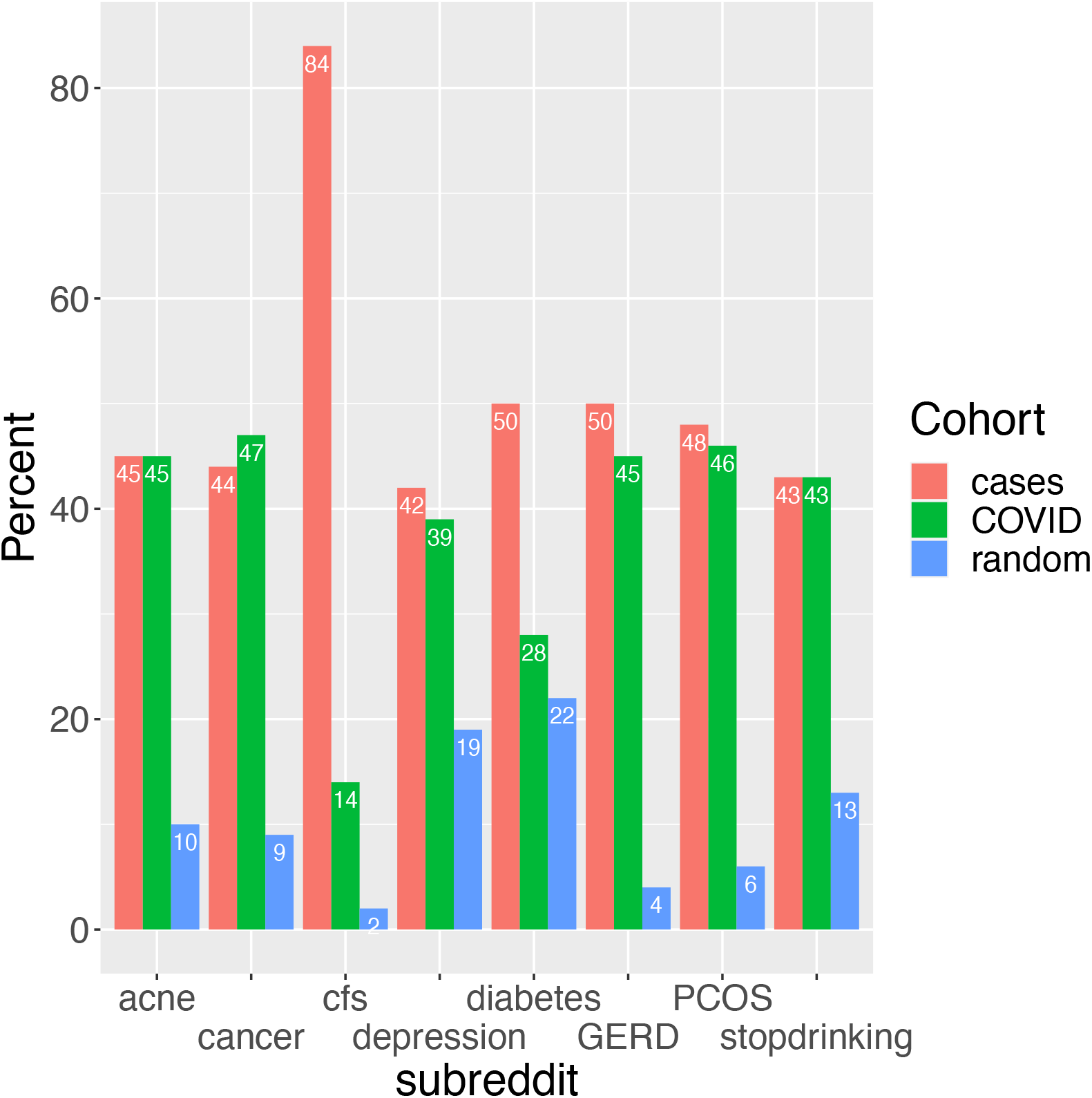
Comparison of r/cfs enrichment among cases, versus enrichment of other medical support forums. For r/cfs and 7 comparison illness support forums, the percentage of study users who posted in the subreddit prior to 2020 by study group. Study groups include cases (red): users who posted to r/covidlonghaulers, COVID (green): users who posted to r/COVID19positive, and random (blue): a random sample of recently active Reddit users.

### Dual posting vs transitioning

We next investigated the extent to which the pre-20202 r/cfs users active on r/covidlonghaulers continued to post on r/cfs after their first post on r/covidlonghaulers. We restricted analysis to the 83 such users for whom we had at least 3 months of follow-up represented among the bulk Reddit archive. Of these 83 users, 53 (64%) continued to post at least once to r/cfs during the follow-up period after their first post on r/covidlonghaulers and the other 30 (36%) did not.

### Qualitative analysis

To more deeply understand the interactions between the r/cfs and r/covidlonghaulers communities, we performed a qualitative analysis of posts written on r/covidlonghaulers by users who were active on r/cfs prior to 2020. We hypothesized that members of the r/cfs community would be driven by a variety of motives to become active on r/covidlonghaulers. Posts were manually reviewed for motives and coded according to rules described in the Methods. We found that motives of r/cfs users active on r/covidlonghaulers included seeking mutual support between communities with some shared experiences, professional interest in illnesses associated with chronic fatigue, and dual identification with both syndromes. Specifically of the 160 pre-2020 r/cfs users later active on r/covidlonghaulers, 54 sought or offered mutual support with long COVID patients as a ME/CFS patient or patient of another chronic illness, 5 stated or implied professional interests related to one or both syndromes, 39 self-identified with both syndromes, and 82 could not be classified into any of these categories (Figure 3). A striking finding among these qualitative findings is the plurality of r/cfs members on r/covidlonghaulers who are there seeking or offering mutual support with long COVID patients as ME/CFS patients. Within the dual identification category, users frequently mentioned that their symptoms after COVID were new, different, or worse than pre-existing symptoms, or that they had been experiencing a significant period of relief from pre-existing symptoms in the time prior to developing COVID and subsequent long COVID. Within the mutual support category, a commonly expressed sentiment was the hope among ME/CFS-identified users that as long COVID becomes better recognized, that ME/CFS too, in light of some similarities with long COVID, will finally receive the attention and respect it deserved.

**Fig. 3:**
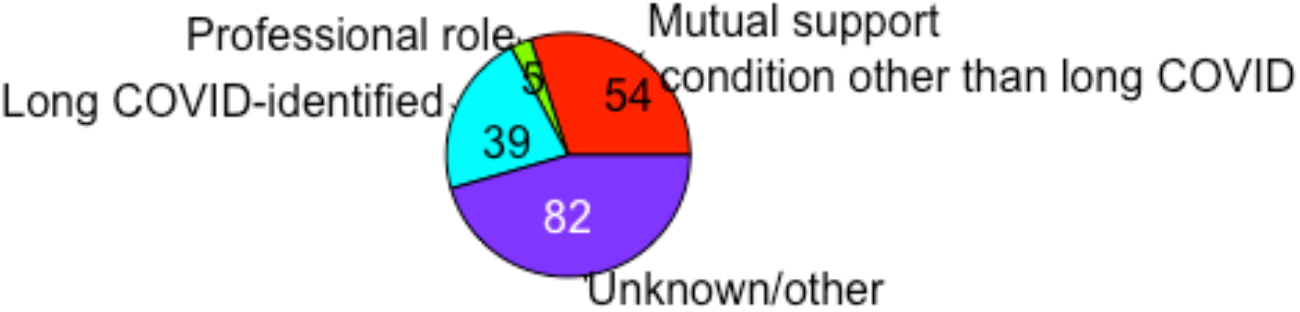
Stated/implied motives of r/cfs users active on r/covidlonghaulers. Pie chart of stated/implied motives of 160 users active pre-pandemic on Reddit’s ME/CFS support forum who later became active on Reddit’s long COVID support forum.

## Discussion

In our retrospective case-control study of Reddit data, we found that users active pre-pandemic on the ME/CFS support forum are exceptionally likely to become active participants on the long COVID support forum. The converse is not true: the vast majority of users active on the long COVID support forum had no prior connection with the ME/CFS support forum. In our qualitative post-analysis review, there were two main types of users on both forums.

The first major type of users on both forums are those with pre-existing ME/CFS symptoms whose symptoms relapsed or progressed after developing COVID-19. One interpretation of this finding is that in these individuals, the long COVID syndrome may represent the same underlying ME/CFS disease, in which COVID-19 plays a role in disease progression. A related interpretation is that in these individuals long COVID and ME/CFS are separate illnesses but that these individuals may be predisposed to fatigue-associated illness. A third interpretation is that ME/CFS’s fluctuating course can make it challenging to ascertain which symptoms are pre-existing and which are due to COVID-19 in this subset of users. Future research to distinguish among these possibilities can shed more light on the underlying nature of both long COVID and ME/CFS.

The second major type of user active on both forums identifies with ME/CFS but not long COVID and are active on the long COVID forum to seek and offer support with patients suffering from a condition with some clinical and social similarities to their own. The high rate of ME/CFS patients seeking mutual support on a long COVID forum speaks to the longsuffering experience of these patients not feeling heard or respected in some segments of society and the medical community.

A strength of this study is its use of mixed methods to more deeply understand its quantitative findings. Limitations of the study include that its sample of Reddit users may not generalize to the larger population, and that users self-reported conditions may not equate with clinical diagnosis. In addition, demographic information about the users is sparse and does not capture the known female-predominance of long COVID (19) or the previously observed but now receding male predominance of Reddit (20).

Overall, these results highlight clinical and social reasons that bring ME/CFS patients to a long COVID forum. Clinically, our results highlight the possibility that patients with pre-existing ME/CFS may be at increased risk of long COVID and/or relapses in ME/CFS due to SARS-CoV-2 infection. This exploratory possibility could be tested in a future study using more traditional clinical data types such as clinical registries or electronic health records. Socially, our results highlight a hope repeated by many of these users that as long COVID becomes better recognized, that ME/CFS too, in light of having some similarities to long COVID, will finally receive the attention and respect it deserves.

## Data Availability

This study produced no new data. The original data from this study were obtained from https://files.pushshift.io.

## Declarations

### Data access

At time of manuscript preparation, raw Reddit data files were obtained from (12)

## Acknowledgments

The authors wish to acknowledge ZB for reviewing a version of the manuscript and providing a patient-perspective. No financial compensation was provided.

## Appendix A Regular expressions for extracting age

~~~
patterns = c(“I am [1-9][0-9]”,
    “I am a [1-9][0-9]”,
    “I \\[[1-9][0-9]”,
    “I \\([1-9][0-9]”,
    “I\\[[1-9][0-9]”,
    “I\\([1-9][0-9]”,
    “me \\[[1-9][0-9]”,
    “me \\([1-9][0-9]”,
    “me\\[[1-9][0-9]”,
    “me\\([1-9][0-9]”)
not_patterns = c(paste0(“[0-9][0-9]”, c(“[0-9]”, “day”, “week”, “wk”, “month”, “hour”, “minute”, “second”, “times”,
            “hundred”, “thousand”, “million”, “billion”, “trillion”,
            “att”, “str”, “combat”, “hit”, “deff”, “summon”,
            “team”, “-something”, “something”, “shades”, “point”,
            “dollar”, “cent”, “\u20ac”, “lb”, “kg”, “pound”, “kilo”,” inch”, “cm”, “feet”, “mile”,
“km”,
            “block”, “house”,
            “page”, “line”, “mg”,
            “%”, “percent”, “\\.”,”‘“,”-petite”, “petite”, “dpo”, “large”, “XL”, “medium”, “small”,
“XS”,
            “level”, “lvl”, “\\+“, “\\+“, “DD”,
            letters[which(!letters %in% c(“f”, “m”))])))
not_patterns = c(not_patterns,”When I am [0-9][0-9]”, “assume I am [0-9][0-9]”, “think I am [0-9][0-9]”, “guess I am [0-9][0-9]”,
     “predict I am [0-9][0-9]”, “believe I am [0-9][0-9]”, “pretend I am [0-9][0-9]”, “imagine I am [0-9][0-9]”,
    “claim I am [0-9][0-9]”, “like I am [0-9][0-9]”, “feel I am [0-9][0-9]”,
    “I am a [0-9][0-9]\\.”, “[0-9][0-9]/[0-9]”, “[0-9][0-9]-[0-9]”)
not_patterns = c(not_patterns, c(paste0(letters, “I \\[[1-9][0-9]”),
            paste0(letters, “I \\([1-9][0-9]”),
            paste0(letters, “I\\[[1-9][0-9]”),
            paste0(letters, “I\\([1-9][0-9]”)))
patterns_extract = c(“.*I am ([1-9][0-9]).*”,
     “.*I ama ([1-9][0-9]).*”,
     “.*I \\[([1-9][0-9]).*”,
     “.*I \\(([1-9][0-9]).*”,
     “.*I\\[([1-9][0-9]).*”,
     “.*I\\(([1-9][0-9]).*”,
     “.* me \\[([1-9][0-9]).*”,
     “.* me \\(([1-9][0-9]).*”,
     “.* me\\[([1-9][0-9]).*”,
     “.* me\\(([1-9][0-9]).*”)
~~~

